# Adaptable Automated Interpretation of Rapid Diagnostic Tests Using Few-Shot Learning

**DOI:** 10.1101/2021.06.23.21258927

**Authors:** Siddarth Arumugam, Jiawei Ma, Uzay Macar, Guangxing Han, Kathrine McAulay, Darrell Ingram, Alex Ying, David A. M. Colburn, Robert Stanciu, Thomas Grys, Shih-Fu Chang, Samuel K. Sia

**Affiliations:** Department of Biomedical Engineering, Columbia University, New York, NY 10027, USA; Department of Computer Science, Columbia University, New York, NY 10027, USA; Department of Electrical Engineering, Columbia University, New York, NY 10027, USA; Department of Laboratory Medicine and Pathology, Mayo Clinic, Phoenix, AZ 85054, USA; Safe Health Systems, Inc., Los Angeles, CA 90036, USA

**Keywords:** Few-shot adaptation, rapid diagnostic tests, self-supervision, COVID-19, pandemic

## Abstract

Point-of-care lateral-flow assays (LFAs) are becomingly increasingly prevalent for diagnosing individual patient disease status and surveying population disease prevalence in a timely, scalable, and cost-effective manner, but a central challenge is to assure correct assay operation and results interpretation as the assays are manually performed in decentralized settings. A smartphone-based software can automate interpretation of an LFA kit, but such algorithms typically require a very large number of images of assays tested with validated specimens, which is challenging to collect for different assay kits, especially for those released during a pandemic. Here, we present an approach – AutoAdapt LFA – that uses few-shot learning, an approach used in other applications such as computer vision and robotics, for accurate and automated interpretation of LFA kits that requires a small number of validated images for training. The approach consists of three components: extraction of membrane and zone areas from an image of the LFA kit, a self-supervised encoder that employs a feature extractor trained with edge-filtered patterns, and few-shot adaptation that enables generalization to new kits using limited validated images. From a base model pre-trained on a commercial LFA kit, we demonstrated the ability of adapted models to interpret results from five new COVID-19 LFA kits (three detecting antigens for diagnosing active infection, and two detecting antibodies for diagnosing past infection). Specifically, using just 10 to 20 images of each new kit, we achieved accuracies of 99% to 100% for each kit. The server-hosted algorithm has an execution time of approximately 4 seconds, which can potentially enable quality assurance and linkage to care for users operating new LFAs in decentralized settings.

## Introduction

Lateral-flow assays (LFAs) present an increasing opportunity for increasing accessibility to diagnostic assays, but errors in assay operation and results interpretation hinder their deployment in decentralized settings, such as primary care clinics and homes^1-5^. For example, improper assay operation could produce absent control bands; alternatively, failure to identify the presence of faint bands or confusing the zone type (e.g., control vs. test bands) could lead to an incorrect interpretation of overall assay result^6^. For example, in the COVID-19 pandemic, a number of SARS-CoV-2 antigen tests are now approved for home use^7^ to support decentralized testing; consequently, user errors from incorrect operation and results interpretation are also likely to become more prominent in the coming months to years.

Image-processing algorithms to automate the interpretation of LFAs and rapid diagnostic tests can potentially provide quality assurance to users in decentralized settings and reduce incidence of these errors, but existing algorithms have shortcomings such as the need to use images collected by smartphones with physical attachments^5,8-14^, are designed to work retrospectively with a library of pre-collected images^15,16^, or require a large number of labelled training images ranging in the hundreds^17,18^ for each assay kit. By comparison, an ideal algorithm would be scalable in deployment (for example, working in real time with images collected by a smartphone camera without an extra adapter) and would not require experimental collection of a large number of expert-labelled images with validated clinical specimens across a large and ever-changing roster of new LFAs, which is especially challenging to achieve during public health emergencies.

In this study, we have developed 1) an end-to-end modular workflow to work with assay kit images taken with a smartphone with no external adapters and 2) a trained algorithm that can be adapted to a new assay kit with just 10 to 20 images of the new kit in order to accurately interpret the result (**Fig. 1a**). The ideal algorithm should generalize to different LFA kits with variations in color and size of bands^1,19-22^: the color and intensity of bands depend on sizes and shapes of gold nanoparticles^20,23^, material properties of the membrane, and membrane pretreatment steps^20,21^; the sizes of bands depend on liquid-dispensing conditions and capillary flow time^20,21^. Here, we propose a few-shot adaptation strategy – which has been employed in applications ranging from computer vision to robotics ^24-26^ to learn a strong classifier for new domains^27-31^ – to mitigate the performance drop caused by domain shift stemming from image-pattern variance, using only a few labeled images. In our few-shot model adaptation strategy, zone areas from many images of a “ base” kit are cropped and used to pre-train a feature-extraction network that employs self-supervised learning with edge-filtered images. To adapt to a new assay kit, zone areas from 10 to 20 images are cropped, and the pre-trained model from the base model (shown as “ B” in **Fig. 1a**) adjusts its weights to a model adapted to a new assay (shown as “ N” in **Fig. 1a**) using supervised contrastive learning. Thus, for the end user operating a new assay kit, zone areas from an image of the kit are cropped, and the adapted network automatically and accurately interprets the bands at each zone and overall assay result.

**Figure 1.**
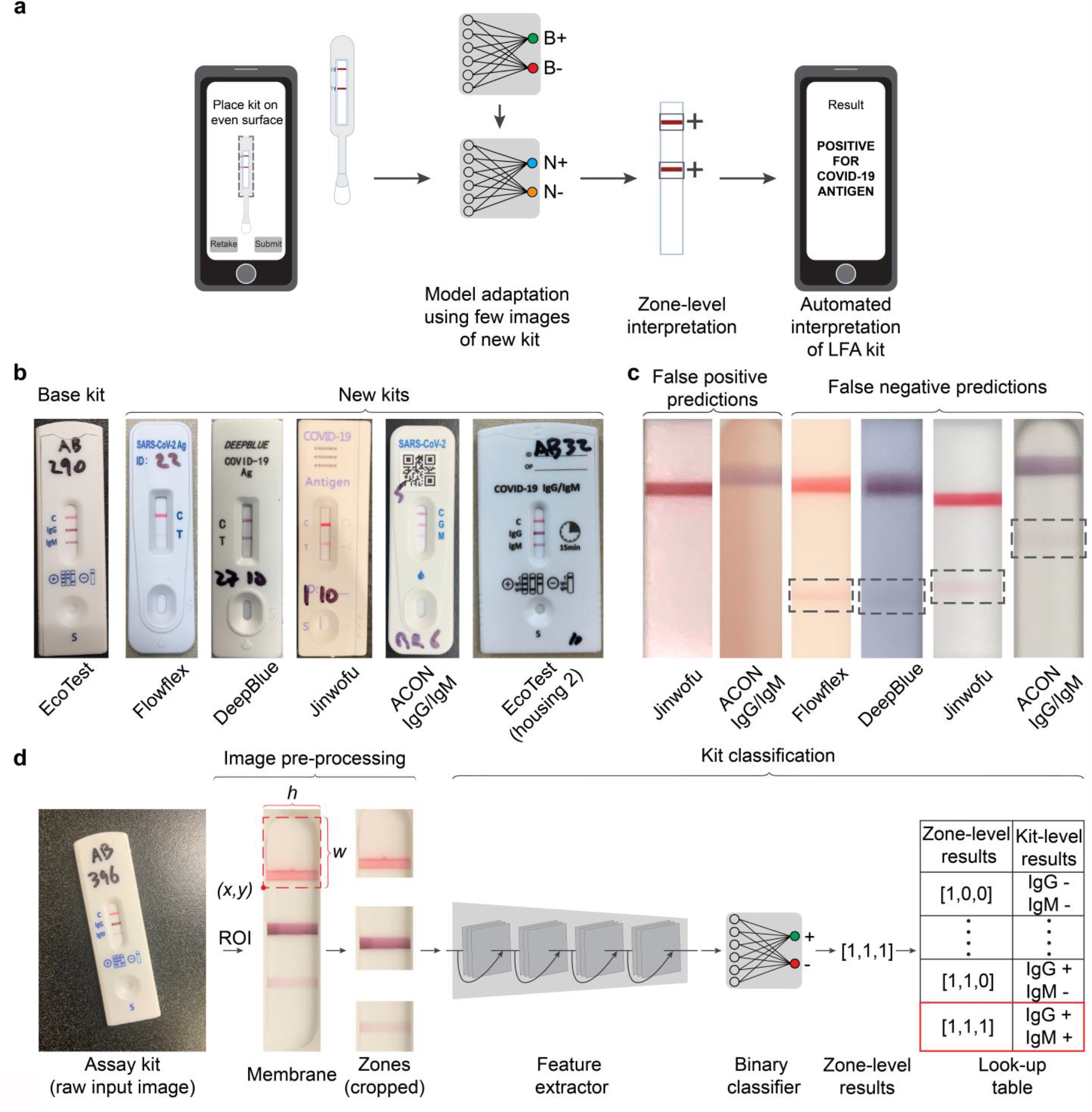
Overview of use case, challenge, and pipeline for image processing and machine learning. **(a)** Envisioned testing process for end user. The user takes an image of the assay kit using a smartphone displaying an on-screen image guide. The zones containing the bands are automatically identified, and a model that was pre-trained on a base kit (shown as “ B” in the network) and previously rapidly adapted to a new kit (using few-shot learning, shown as “ N” in the network) processes the images of zones. The model classifies each zone as positive or negative, and provides an overall assay result on the screen of the smartphone. The cloud-hosted model processes the image and generates the results in ∼4 seconds. **(b)** Images of a base LFA kit (EcoTest) for pre-training the model, and five new COVID-19 LFA kits (including both antigen and antibody tests) to be interpreted. **(c)** Images illustrating the challenge for few-shot learning. A pre-trained model on the base kit, without adaptation, produces failed predictions on the new kits. Shown are both false positives and false negatives (likely due to variations in colors and intensities of membrane background and bands). **(d)** Overview of AutoAdapt LFA pipeline. From a raw input image of an assay kit, a correction of orientation and perspective is applied to segment an image of an assay kit. From the assay kit image, a segmentation model based on Mask R-CNN is used to extract the membrane region of interest (RoI). Based on measured kit-specific parameters, individual zones are cropped, and passed through a software pipeline consisting of a feature extractor followed by a binary classifier. Classification of each zone allows, via a kit-specific lookup table, for a final classification of assay result (“ kit-level” classification or result) as positive, negative, or invalid.

In this study, we pre-trained a base model using expert-labelled images from the AssureTech EcoTest COVID-19 IgG/IgM Antibody Test, an assay authorized by FDA, and adapted the model to interpret LFAs from five other commercial COVID-19 LFAs (**Fig. 1b**). The five LFAs include three antigen tests (ACON Flowflex SARS-CoV-2 Antigen Rapid Test, Anhui DeepBlue SARS-CoV-2 Antigen Test, and Jinwofu SARS-CoV-2 Antigen Rapid Test), one antibody test (ACON SARS-CoV-2 IgG/IgM Antibody Test), and an AssureTech EcoTest COVID-19 IgG/IgM Antibody Test kit that uses a different housing (denoted in the paper as ‘EcoTest (housing 2)’) but retains use of the same LFA membrane (**Fig. 1b**). Like almost all commercial LFAs, these kits share rectangular control and test bands, but differ in kit housing dimensions and membrane dimensions, as well as number, spacing and color of bands (kit-specific dimensions shown in **Supplementary Table 1**). To illustrate the challenge for interpreting new assay kits, without few-shot adaptation, this pre-trained algorithm produced incorrect predictions (both false positives and false negatives) due to variations in color and intensities of bands and membranes across new LFA kits (**Fig. 1c**).

### Overview of pipeline

In the AutoAdapt LFA algorithm (**Fig. 1d**), a user takes an image of an LFA kit, which enters a cloud-hosted pipeline with an instance-segmentation model that corrects the orientation and perspective of the raw image, segments the assay kit from background and the membrane from assay kit, and crops individual zones (i.e., regions in the membrane corresponding to bands and a portion of surrounding area) from the membrane. Next, images of zones enter a feature-extraction network, which is learned in order to generate robust feature representation as unique signatures to discriminate positive cases from negative cases under diverse conditions (e.g., color, intensity, and width of bands); the feature-extraction network also adapts to new LFA kits with a small number of images. From latent feature vectors for each zone, a binary classifier recognizes colored rectangular bands, the form factor seen in the vast majority of LFAs^32,33^, and determines whether a band is present or absent in each zone. Finally, an assessment of the LFA kit result is obtained by comparing the output of the binary classifier with a lookup table containing all combinations of possible zone-level classification results; this kit-level classification is sent to the user’s smartphone as an interpreted LFA result. The server-hosted algorithm has a mean execution time of 3.55 ± 2.28 seconds. Overall, this rapid automated interpretation pipeline could fit within a larger digital platform^34-37^ that collects demographic data for epidemiology and provides instructions to carry out the test as well as follow-up linkage to care; **Supplementary Fig. 1**.

To develop this pipeline, we framed the objective as learning the optimal parameters of the feature-extraction network and the classifier module by minimizing the loss functions given a set of training images for an assay kit. Unlike methods that require *de novo* training on a new LFA kit, we developed two novel methods to achieve adaptation requiring only a small number of images of new kits. First, to ensure the underlying feature representation is robust against variations in the LFA images, we developed a feature extractor that learns to extract robust latent representation of zone images for classification; these latent representations are also used to reconstruct (decode) the edges associated with the images. This auxiliary edge reconstruction task is in addition to the standard fully-supervised classification task and helps learn feature representation for effective adaptation, which is based on the observation that edges in an image tend to remain invariant in diverse LFA images. As shown in **Fig. 2a**, edge-preservation can be learned in a self-supervised manner (not requiring manually-assigned labels), by using the output of an automatic edge detection algorithm (Sobel filter^38^) as the ground truth for decoding. Second, as shown in **Fig. 2b**, our system performs supervised contrastive learning using a dataset combining images from the new LFA kit and the base LFA kit and learns a new classifier for the new kit. Here, the neural network model pre-trained on a set of labeled LFA images from the base kit is adapted to a new target LFA, using 10 to 20 labelled training images of a new kit.

**Figure 2.**
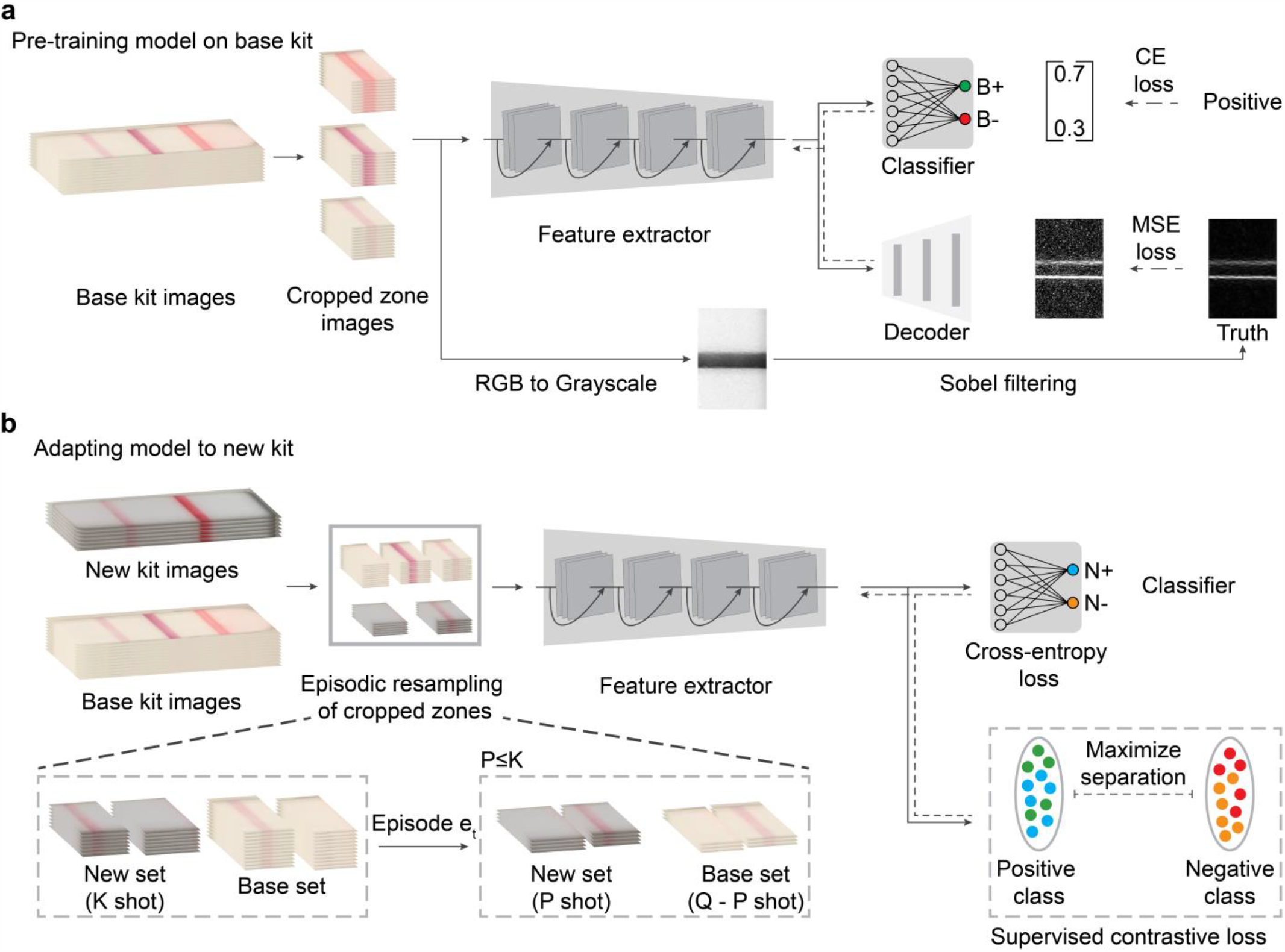
Self-supervision and few-shot adaptation for LFA kits. **(a)** The feature extractor is pretrained on the base kit using self-supervised learning task over edge-filtered patterns and fully-supervised binary classification task. For each zone, fully-supervised binary classification is carried out with cross-entropy loss with the annotated binary labels. Sobel filter is used to highlight the edge pixels between the band and the background of the membrane. The edge image after normalization is used as ground truth and the learning process is used to reconstruct an image that resembles the ground truth edge image, with the quality measured in MSE (Mean Square Error). The solid and dashed arrows indicate forward processing and gradient backpropagation respectively during the learning process. **(b)** Model adaptation is carried out by supervised contrastive learning to regularize the feature extractor and fully-supervised learning to learn an adapted classifier for the new kit. A sampling strategy to build an episode with Q (e.g., 32) images per class is used: for each class (positive or negative), given K (e.g., 10) images available, P (e.g., 4) images are subsampled from the new kit and mixed with Q-P images of the base kit.

### Image pre-processing

For model training as well as during inference, the first module corrected for skew and extracted the zones from the images of LFA kit. This module first detected the orientation of the kit and carried out perspective correction using the predicted segmentation mask of the LFA kit (**Supplementary Fig. 2**). This mask was generated by using Mask R-CNN^39^, an instance segmentation model (more details in Methods). The kit membrane from the perspective corrected image was then localized and individual test zones were cropped out using the kit-specific dimensions listed in a JSON file. For this study, the test-specific dimensions, such as kit height, kit width, membrane width, membrane height, and zone dimensions, were measured from images of LFA kits using Adobe Photoshop v21.0.2 and saved as a JSON file. These dimensions could be directly provided by the kit manufacturers in the future. To measure the accuracy of the automatic membrane segmentation step, we measure the intersection over union (IoU) scores between the segmented membrane and the manually annotated ground-truth membrane region. IoU scores greater than 90% for all the assay kits (**Table 1**) confirm the robustness of this first step.

**Table 1.** Intersection over Union scores for membrane segmentation. The IOU scores for each of the new kit images was obtained by selecting ten images at random from a labelled pool of 30 images for training and evaluating the performance on a fixed evaluation set of ten images.

| Kit name | IOU score |
| --- | --- |
| Flowflex | 0.92 |
| DeepBlue | 0.89 |
| Jinwofu | 0.90 |
| ACON IgG/IgM | 0.93 |
| EcoTest housing 2 | 0.93 |

### Pre-training of feature extractor with edge detection and self-supervised learning

The cropped test zones were fed into a feature extractor and the extracted features were passed into a binary classifier and a decoder (**Fig. 2a**). The binary classifier (a fully connected layer) outputs ‘0’ or ‘1’ to denote the absence or presence of the band in the cropped zone, respectively. The images from the base LFA kit were manually annotated with the binary labels, and the classifier was trained to learn specific prototypes associated with the positive and negative classes using cross-entropy (CE) loss.

Images of kits with faint bands can lead to false negatives while stained membranes and lighting artifacts can lead to false positives (**Fig. 1c**). Even though such failure cases can be reduced by training on a large number of relevant examples, acquiring sufficient images on a new LFA kit present a logistical challenge. Directly applying the model conventionally trained on the base LFA kit (i.e., by minimizing only the CE loss) to new LFA kits, resulted in low classification accuracy on the new kits. Hence, we designed a self-supervised, edge-enhanced image reconstruction task to improve the generalizability of the feature extractor (**Fig. 2a**). The network was trained to detect the edges of the image pattern (pixels at the junction between the membrane background and the band in the zone) and reconstruct the corresponding edge-enhanced image. This task is self-supervised: starting with RGB images of zones from a base LFA kit, the model converted the image into grayscale and applied a Sobel filter^38^ to generate the ground truth image set (Sobel filter is a basic image processing algorithm that generates an image emphasizing edges). In parallel, the model fed the extracted features into the decoder to reconstruct the edge-enhanced image. This model was then trained to minimize the mean squared error (MSE) between the reconstructed edge image from the decoder and the ground truth edge image. By combining the fully-supervised image classification with the self-supervised edge-enhanced image reconstruction, the feature extractor, classifier, and decoder were trained jointly to optimize the zone classification accuracy as well as to learn a good feature representation that is sensitive to edge information.

### Learning a new classifier for a new LFA using few-shot adaptation

The pre-trained model from a base LFA kit was adapted to a new LFA kit with minimal retraining via few-shot adaptation (**Fig. 2b**). We mixed the labeled data of the base LFA kit and the new LFA kit and used this as the training set. We specifically used this mixture of data from new kit and base kit to avoid overfitting to the small number of images of the new kit. In addition to the CE loss used to train the binary classifier for the new LFA kit, we also used supervised contrastive learning, between the cropped zone images of both the new kit and the base kit, to refine the feature extractor.

We gathered the cropped zone images of both the base kit and the new kit, resampled the data, and calculated the supervised contrastive (SupCT) loss^40^. First, we extracted features of the base kit cropped zone images for both positive and negative classes and considered them as anchors. Next, we extracted features from the cropped zone images of the new kit and compared them with all of the anchors using cosine similarity. The feature extractor was then trained to maximize the cosine similarity between features of the same class. For the implementation, we resampled the cropped zone images from the mixed dataset to build episodes and then computed SupCT loss within each episode (more details in Methods). As a comparison to the adaptation strategy, we also performed fine-tuning which only calculated the CE loss among samples within the episodes for network updating.

## Results

### Description of datasets

While gathering the image dataset, we varied imaging conditions by using different phones and imaging the assay kits under varied lighting conditions (more details in Methods). For pre-training of the model, the training dataset from the base kit (AssureTech EcoTest COVID-19 IgG/IgM Antibody Test) consisted of 383 membrane images (674 positive zones and 475 negative zones). An additional 254 membrane images (441 positive zones and 254 negative zones) were used as the validation set for model selection under the fully-supervised classification task.

In addition, we used a variational autoencoder^41^ to generate a synthetic dataset composed of 600 zones each of faint positive and negative zones^42^. The synthetic data was mixed with the training dataset for the self-supervised edge-reconstruction task.

The performance of base model is reported on an evaluation set consisting of 102 membrane images (168 positive zones and 138 negative zones) of the base kit. The results (**Table 2**) demonstrate that our model works well for both zone-level classification and overall kit-level classification on the base kit. A “ zone-level” classification accuracy is the model’s performance on all the zones for the entire evaluation data set, and “ kit-level” classification accuracy is the model’s performance in classifying all constituent zones of a single kit (e.g., a kit-level result would be incorrect if any zone in that kit was classified incorrectly). Details regarding the dataset for the five new kits are provided in **Supplementary Table 2**.

**Table 2.** Zone-level and kit-level classification accuracy without adaptation (direct testing) and with adaptation. For the direct testing case, the model pretrained on the base kit was directly applied on each of the new kit’s evaluation dataset. For the adaptation approach, the pretrained model was adapted to each of the new kits, except for EcoTest housing 2 kit, using 10-shot adaptation (20 zone images) and the performance on their respective evaluation datasets is listed here.

| Kit name | Without adaptation |  | With adaptation |  |
| --- | --- | --- | --- | --- |
|  | Zone accuracy (%) | Kit accuracy (%) | Zone accuracy (%) | Kit accuracy (%) |
| EcoTest (base) | 98.8 | 96.5 | - | - |
| Flowflex | 93.1 | 86.1 | 99.8 | 99.6 |
| DeepBlue | 93.2 | 86.4 | 99.5 | 98.9 |
| Jinwofu | 94.7 | 89.4 | 100 | 100 |
| ACON IgG/IgM | 91.0 | 73.6 | 98.8 | 96.4 |
| EcoTest housing 2 | 100 | 100 | - | - |

### Performance on 5 new COVID-19 tests

We employed the pre-trained feature extractor using the few-shot adaptation strategy on five COVID-19 LFA kits, and assessed the effects of our adaptation strategy and self-supervised edge-detection task separately. The performance of the base model on the new kits are shown when applied directly and with the proposed adaptation method using 10-shots (20 zone images) highlighting the significant performance improvement seen using our few-shot adaptation strategy (**Table 2**). On top of the pretrained base model, adaptation can consistently improve the performance by including only a few training images of the new LFA kits. The EcoTest housing 2 kit was identical in all aspects to the base kit expect for the housing, so the direct application of the base model without any adaptation was able to achieve 100% zone-level and kit-level accuracies.

In **Fig. 3**, we plot the classification accuracy, at zone level and kit level, against the number of zone images used during the adaptation process, ranging from 0 (direct testing) to using the entire training dataset. These figures also serve as the ablation study evaluating the separate contributions made by self-supervision in pretraining the feature extractor as well as the supervised contrastive learning during adaptation. We compare our adaptation approach with three alternative approaches: 1) the proposed approach without the self-supervision component in the pre-training stage, 2) the proposed approach without supervised contrastive loss during adaptation, and 3) training the network for a new kit from scratch without the two components. The second approach can be considered as a finetuning process that uses the pre-trained base model and finetunes it with the standard CE loss. For all approaches, the base kit and new kit images were mixed for network training, and the same data sampling strategy was used to ensure a fair comparison.

**Figure 3.**
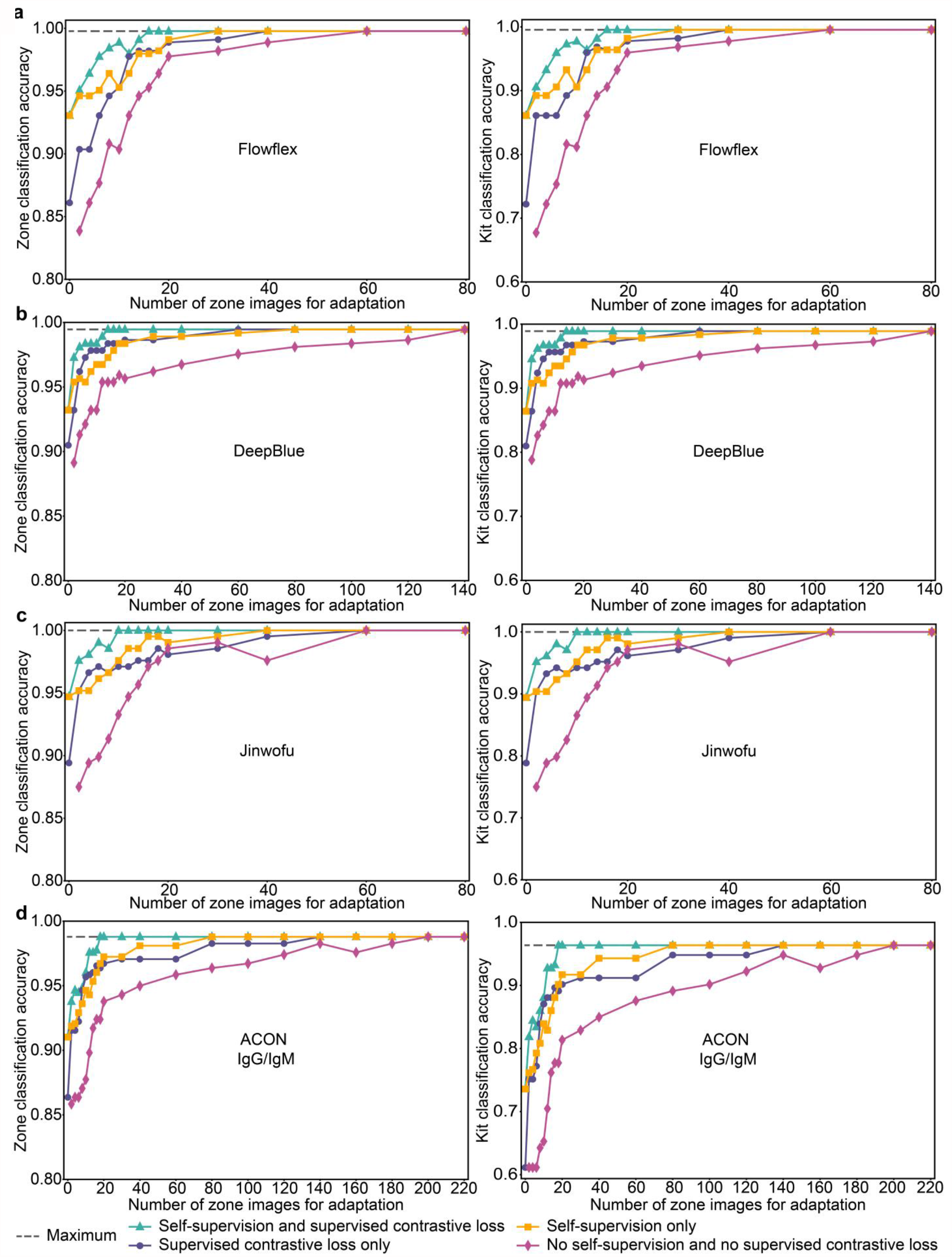
Zone-level and kit-level classification accuracies for four new COVID-19 LFA kits shown with ablated models and number of new kit training images. Ablation studies were carried out to analyze the relative contributions of self-supervised learning for feature extraction and supervised contrastive learning for adaptation. Each model was evaluated by varying the number of images used in the adaptation. Zone-level accuracy scores (left) and kit-level accuracy scores (right) reported for four new assay kits, **(a)** Flowflex, **(b)** DeepBlue, **(c)** Jinwofu and **(d)** ACON IgG/IgM. (The EcoTest housing 2 kit was identical in all aspects to the base kit expect for the housing, so the direct application of the base model without any adaptation was able to achieve 100% zone-level and kit-level accuracies.) The maximum accuracy indicates the upper bound attained by training a model from scratch using all training images for each kit.

For each kit, a random set of images of each class were selected from the training dataset for model adaptation and performance of the trained model was validated against a separate evaluation dataset. The plots for the different approaches are compared against the performance upper bound achieved when using all the new images available for training mixed with the base training images for the classification and edge reconstruction tasks. We showed that for each of the kits, Flowflex, DeepBlue, Jinwofu, and ACON IgG/IgM we achieved maximum classification accuracy using just 16, 14, 10, and 18 zone images respectively for the adaptation. For example, we were able to adapt the base model to the Flowflex kit (**Fig. 3a**) using only eight zone images per class (16 zone images) and reach the same performance (99.8% and 99.6% for the zone and kit levels respectively) as a model trained from scratch using all available training data (200 zone images). The results confirm that both self-supervised pretraining and supervised contrastive loss help, and the combination of these two key ideas helps reach the highest attainable performance. Between these two novel ideas, supervised contrastive learning is more effective: it requires fewer training images during adaptation in order to reach the performance upper bound that is achieved by using the entire training dataset.

In addition, as the feature extractor is pretrained under self-supervision, the extracted features are sensitive to the edges and can work well even when zones with faint bands are encountered. Even though the ACON IgG/IgM kit had the highest frequency of faint bands in our dataset, our approach was able to reach the same performance as using entire training dataset (**Fig. 3d**) using only nine images of each class (18 zone images). Adaptation without supervised contrastive learning can also reach the same performance using 40-shot adaptation. For the model trained without self-supervised pretraining 70 images (with SupCT loss) and 100 images (without SupCT loss) of each class were required to reach the best performance. In addition, direct testing performance (0-shot adaptation) of the model pretrained on the base kit was higher when trained using self-supervision than when trained using only the CE loss.

**Table 3** shows the confusion matrices of the performance of the optimum shot adaptation when evaluated on the evaluation dataset. By starting with a base model pretrained on an existing LFA kit (AssureTech EcoTest COVID-19 IgG/IgM antibody assay kit), we have shown that it is possible to adapt the existing model to different assay kits, which have different numbers of test lines and form factors, using a small fraction of the images needed to train the base model with no loss in accuracy. In addition to evaluating the confusion matrix among on samples in the evaluation set, we devised an ambiguity region to evaluate the distribution of detection scores (probability of positive class). The ambiguity region is bounded by the detection score thresholds such that an image will be correctly classified only if the probability of the ground truth class is high. The thresholds can be either manually set or statistically estimated with 95% area under the curve (more details in Methods). We checked the detection scores of all the images in the evaluation dataset against the ambiguity regions and those images with scores falling in ambiguity region were not classified. We computed the percentage of images that were categorized as ambiguous as well as the accuracy over the images that were classified. Since the detection score for the false predictions were close to 0.5, they fell into the ambiguity region. Therefore, by using this concept of the ambiguity region we were able to treat most of the failure cases as ambiguous while keeping the number of true predictions that fell into the ambiguity region to a minimum. This further increased the classification accuracy among the classified samples consistently over four new target kits (**Table 3**).

**Table 3.** Confusion matrices of the best models applied on the evaluation dataset (left) Accuracy and the percentage of ambiguous kits across varying ambiguity regions (right) **(a)** is the confusion matrix for the base model on base kit evaluation set. The performance of the best performing adapted model on the evaluation dataset for each of the new kits **(b – e)** without the enforcement of an ambiguity region is shown in the tables to the left. (**f)** shows the confusion matrix for the EcoTest housing 2 kit. The tables on the right show the accuracy for the corresponding assay kit and percentage of kits classified as ambiguous values for the different kits when varying the ambiguity region.

**a) EcoTest (base kit)**
| Prediction \ Label | Positive | Negative |
| --- | --- | --- |
| Positive | (TP) 165 | (FP) 1 |
| Negative | (FN) 3 | (TN) 137 |

**b) Flowflex (2-zone kit; 224 zone images)**
| Prediction \ Label | Positive | Negative | Ambiguity Region | [0.50, 0.50] | [0.08, 0.98] | [0.20, 0.80] |
| --- | --- | --- | --- | --- | --- | --- |
| Positive | (TP) 303 | (FP) 0 | Accuracy | 99.8% | 100% | 100% |
| Negative | (FN) 1 | (TN) 142 | Percentage ambiguous kits | - | 5.2% | 1.3% |

**c) DeepBlue (2-zone kits; 184 zone images)**
| Prediction \ Label | Positive | Negative | Ambiguity Region | [0.50, 0.50] | [0.21, 0.95] | [0.20, 0.80] |
| --- | --- | --- | --- | --- | --- | --- |
| Positive | (TP) 242 | (FP) 0 | Accuracy | 99.5% | 100% | 100% |
| Negative | (FN) 2 | (TN) 124 | Percentage ambiguous kits | - | 4.9% | 4.1% |

**d) Jinwofu (2-zone kits; 104 zone image)**
| Prediction \ Label | Positive | Negative | Ambiguity Region | [0.50, 0.50] | [0.05, 0.99] | [0.20, 0.80] |
| --- | --- | --- | --- | --- | --- | --- |
| Positive | (TP) 164 | (FP) 0 | Accuracy | 100% | 100% | 100% |
| Negative | (FN) 0 | (TN) 44 | Percentage ambiguous kits | - | 1.4% | 0 |

**e) ACON IgG/IgM (3-zone kits; 193 zone images)**
| Label<br>Prediction | Positive | Negative |
| --- | --- | --- |
| Positive | (TP) 386 | (FP) 1 |
| Negative | (FN) 6 | (TN) 192 |

| Ambiguity<br>Region | [0.50,<br>0.50] | [0.11,<br>0.88] | [0.20,<br>0.80] |
| --- | --- | --- | --- |
| Accuracy | 98.8% | 99.5% | 99.3% |
| Percentage<br>ambiguous<br>kits | - | 3.5% | 2.8% |

**Table 3. Confusion matrices of the best models applied on the evaluation dataset (left)**
| Label<br>Prediction | Positive | Negative |
| --- | --- | --- |
| Positive | (TP) 66 | (FP) 0 |
| Negative | (FN) 0 | (TN) 6 |

### Conclusions and future work

We have described the development of AutoAdapt LFA, an approach for the adaptation of a LFA kit interpretation model trained on one kit to new kits, each with a different form factor. We showed that this adaptation can be carried out using a much smaller subset of images than what was used for training the base model. Compared to *de novo* training on every new assay kit, this reduction in the number of images was achieved by adopting a modular approach to the machine-learning pipeline: starting from an image of the kit, the perspective-corrected membrane and individual zones were extracted followed by the extraction of the features preserving edge information, and finally a binary output which indicated whether a band was present in the cropped zone. A robust feature extractor is important for handling challenging images in LFA kits like those with faint or partially formed lines. Our approach of using self-supervision to extract features preserving edge information addressed this issue, and it is believed that this use of self-supervised learning to reconstruct edge-enhanced images has not been previously demonstrated. To our knowledge, the application few-shot learning, including this adaptation framework, has not been demonstrated for interpretation of LFA kit images. Thus, we have shown that using this novel approach, we can train accurate classification models using a fraction of kit images that would be required in *de novo* training.

In terms of impact for medicine, this reduction in new training images to achieve assured user interpretation of rapid test images is significant with the rise of use of rapid diagnostic tests. Most immediately, the COVID-19 pandemic has thrusted front and forward the need for rapid testing and population surveillance to track and control the spread of the disease in a scalable and timely manner. If effectively implemented, point-of-care testing can contribute significantly to a rapid and effective public health response – as well as patients’ individual safety, privacy, physical health and mental well-being – by enabling widespread timely testing in a manner that does not overwhelm the limited capacity of testing facilities or provoke social crowding at selected testing sites. By expediting the process of training a model to newly available rapid diagnostic tests, the AutoAdapt LFA approach could facilitate reliable decentralized testing and real-time monitoring of disease prevalence. In the longer term, the need to achieve assured user interpretation will rise as patients and consumers will more frequently monitor their health via self-testing for both infectious diseases and chronic conditions, in an age of precision health. Future work includes validation on a wider variety of rapid tests, and generalization to LFA kits beyond rectangular bands (for example, as in some vertical flow assays) and bands of single colors (for example, some urinalysis kits with color-based readouts).

## Methods

### Dataset Collection

Base kit (AssureTech EcoTest COVID-19 IgG/IgM Antibody Test): train and validation datasets were gathered using iPhone X at the Mayo Clinic Hospital, Phoenix, AZ. The evaluation dataset images were gathered using three phones by two users: iPhone X, iPhone 7, Samsung Galaxy J3 (SM-J337V). Care was taken to ensure that the kits were imaged under three different ambient lighting conditions (warm white, cool white, and daylight).

Novel kits (ACON Flowflex SARS-CoV-2 Antigen Rapid Test, Anhui DeepBlue SARS-CoV-2 Antigen Test, Jinwofu SARS-CoV-2 Antigen Rapid Test, and ACON SARS-CoV-2 IgG/IgM Antibody Test): training and evaluation sets were gathered using iPhone X at the Mayo Clinic Hospital, Phoenix, AZ. Serum samples for the antibody tests were collected under Mayo Clinic IRB 20-004544 or shared by the Department of Laboratory Medicine at the University of Washington School of Medicine (Seattle, WA)^43^. The use of excess clinical specimens was reviewed by the Mayo Clinic Biospecimens Committee and an appropriate Material Transfer Agreement was drawn up to allow access to de-identified specimens from the University of Washington School of Medicine. The University of Washington IRB approved this work with a consent waiver. Nasopharyngeal swabs from Mayo Clinic Hospital patients were heat fixed and run for the antigen tests under Mayo Clinic IRB 20-010688. All necessary patient/participant consent has been obtained and the appropriate institutional forms have been archived. All assay kits were imaged within 10 minutes of running the test.

### Image acquisition and pre-processing based on Mask R-CNN

The image processing workflow starts with an image of the assay kit being taken by the user through the SMARTtest application^18^ in a fixed portrait orientation. This image is saved in an AWS S3 bucket as an JPEG image from the frontend, and the corresponding URL is sent to the AWS Lambda Function. The function reads the image data, stores the original resolution image in a copy, and resizes the image while preserving the aspect ratio by capping the height of the image to a maximum of 800 pixels. The membrane is localized in the resized image using the instance segmentation model Mask RCNN (**Supplementary Fig. 2**), and the predicted bounding box coordinates in the resized image are then transformed to the corresponding coordinates in the image of the original resolution to get the highest possible resolution of the membrane which is then sent to the classifier.

Mask R-CNN^39^ builds on top of the preceding Faster R-CNN^44^ and Fast R-CNN^45^ models and combines them with a fully-convolutional network (FCN) and introduces object mask prediction (i.e., segmentation^46^) in parallel to bounding box regression. Given an input image, the model extracts feature maps via a pretrained deep neural network (e.g., VGG16), and subsequently passes these in parallel through a ROI-specialized pooling layer followed by several fully-connected layers and an FCN. The instance segmentation model has been trained for two object classes: the kit and the membrane. The model outputs i) detection scores, ii) bounding boxes, and iii) segmentation masks of a maximum of 100 objects. The bounding box defines a rectangular area that contains the assay kit or the membrane. The segmentation mask includes all the pixels that correspond to the actual area of the assay kit or the membrane and do not necessarily have to be rectangular in shape. From all the detected objects we retain information for a kit and membrane object with the highest detection score greater than 0.9. **Supplementary Fig. 3** illustrates different IoU scores and the corresponding membrane segmentation masks for the EcoTest (base kit).

The bounding boxes and segmentation masks of the kit and membrane with the highest detection score are retrieved and a binary segmentation mask is generated for both kit and membrane. Next, the rotation angle is estimated by performing contour detection on the segmentation mask of the kit and membrane, and approximating a minimum-area quadrilateral mask whose corner coordinates can be used to construct a right-angle triangle. The membrane is cropped from the input image with the binary segmentation mask, and is subsequently rotated by the estimated angle. The rotated membrane will have black regions if the estimated angle is greater than zero, and the largest rectangle that doesn’t include any black pixels is estimated and extracted as the final membrane to be sent to the classifier. Additionally, we have the capability to compute the homography matrix^47^ between the predicted segmentation mask and bounding box of the kit, and use it to transform the kit of the image to correct for distortion along the pitch axis.

### Pre-training with self-supervised learning

The model uses the Mean Squared Error (MSE) between the decoder output (the reconstructed image) and the ground truth edge-enhanced image as the loss. For the base kit, the number of labeled images were sufficient so that both the classification and the edge-enhanced image reconstruction tasks were carried out to learn a good feature extractor. Thus, as shown in **Fig. 2a** output features of each cropped zone are sent to both the classifier and the decoder. The model uses the cross-entropy (CE) loss for the classification task and uses the MSE between the reconstruction and the automatically reconstructed edge filtered image to learn the optimal convolution kernel in the decoder for the self-supervised edge reconstruction task. By using the edge-enhanced features, the feature extractor was able to generalize well on new assay kit images even if the zones were faint.

To generate the ground-truth of the self-supervision task, the model first converted the RGB image into a grayscale image, and then performed edge filtering using Sobel filtering to highlight the pixels in the edge region (if an edge exists). The edge filtered images are then normalized between 0 and 1 and set as labels for the self-supervision task.

With the annotated classification label and the self-generated edge detection label, the equally weighted CE loss and MSE were summed up and used as the objective. In this manner, the extracted features were made sensitive to the edge region and the encoded edge information was used for the classification of cropped zone images including those with faint bands.

### Hyperparameter Selection

#### Instance segmentation model structure

We used the ResNet50 CNN as the backbone of the Mask R-CNN and pretrained it on the ImageNet1K dataset for model initialization. The backbone has been trained on ImageNet1K as a fully-supervised image classification task among 1,000 classes. We used a hidden layer size of 256 for the mask predictor.

#### Instance segmentation training

We used 50 epochs and Adam optimizer for all of the training processes. We pretrained the model on a training subset of 50 images of the base kit with a learning rate of 5E-5 and achieved an IOU score of 0.93 on an evaluation set of ten images. We then finetuned the model on the new assay kits with a learning rate of 5E-6 using 10 training images and evaluated the performance on 10 evaluation images. We used the following train-time augmentations: (i) horizontal flip, (ii) scaling, (iii) aspect-ratio modification, (iv) brightness adjustment, (v) contrast adjustment, (vi) hue adjustment, (vii) saturation adjustment, (viii) color distortion, (ix) jitter addition, (x) cropping, (xi) padding, and (xii) Gaussian noise addition. **Supplementary Table 3** shows the results from the test of robustness of the instance segmentation model using bootstrapping.

#### Classification model structure

We used the ResNet18 CNN as the feature extractor and pretrained the model on the ImageNet1K dataset for model initialization^48^. The feature extractor has been trained on ImageNet1K as a fully-supervised image classification task among 1,000 classes. As shown in **Fig. 2a**, during the pretraining on base kit images, classifier is configured as a fully connected layer (top output) and the decoder is configured as a stack of three deconvolution layers (bottom output).

#### Classification model pre-training

Given a training set, all the images were fed into the model in sequence and the loss was calculated for both gradient backpropagation and for updating the model. A single epoch is completed when the model has seen all the images once. 90 epochs were run in our training process. The performance of the model on the validation dataset was determined after each epoch and the model achieving the highest accuracy was selected.

#### Classification model adaptation & finetuning

The network was trained for 100 epochs for each of the new kits with a learning rate of Within each epoch, we sample 30 episodes and set Q (number of samples per class) as 32 for each episode. The feature extractor was tuned with a learning rate of 0.0001. Adam optimizer was used for the network parameter update of both the feature extractor and the classifier. The inbuilt PyTorch image transformation functions were used, namely: 1) horizontal flip, 2) Random Rotation, 3) Color Jitter (including grayscale). **Supplementary Table 4** shows the results from the test of robustness of the adapted model on the 4 new test kits using bootstrapping.

#### Classification model training from scratch

Similar to the initialization step before self-supervision, a ResNet18 CNN is used as the feature extractor which has been trained with the ImageNet1K dataset as a fully-supervised image classification task. The network is then trained on the training images of the new assay kit with Adam optimizer and a learning rate of 0.001. The same transformation functions used for the adaptation were used here.

### Threshold Determination and Ambiguity Region

In general, the thresholds (*δ*_*neg*_, *δ*_*pos*_) for negative class and positive class were determined individually by feeding the detection score (probability of positive, *P*_*pos*_) of all images of each class into the statistical model and fitting separately. Using the threshold determination of positive class as an example, the steps are explained below:

1. Select the Inverse Gaussian Distribution as the model template to be fitted^49,50^. The reasons why we select this one-side distribution model are,
  a. The inverse gaussian distribution is used to model variables of non-negative values.
  b. Since the probability output from the model is between 0 and 1, the inverse gaussian distribution is selected as it is tighter within the range [0,1] (i.e., the area under its probability density function (PDF) curve within [0,1] is closer to one), compared to other distribution models such as Gamma distribution which may have an observable tail in [1, infinity) interval.
2. Feed the *P*_*pos*_ of all labelled positive zone images into the statistical model and use the fitted parameters to draw the PDF curve.
3. We set the area under the probability distribution curve (between the threshold and the extreme value, i.e., 1 for positive and 0 for negative) as 95% and use Divide and Conquer to find the corresponding threshold value *δ*, which is threshold for positive class *δ*_*pos*_.

For a negative class, *P*_*pos*_ is still used as input to find the classification score threshold *δ*_*neg*_. For the convenience of presentation, [*δ*_*neg*_, *δ*_*pos*_] is used to denote the ambiguity region where images with *δ*_*neg*_ ≤ *P*_*pos*_ ≤ *δ*_*pos*_ will not be classified since they fall within the region, and the images with *P*_*pos*_ ≤ *δ*_*neg*_ or *P*_*pos*_ ≥ *δ*_*pos*_ are classified as negative or positive respectively. The ratio of the unclassified images with respect to the entire evaluation set is reported as the percentage of ambiguous cases (as shown in **Table 3**).

## Data Availability

The image library and code are available on request.

## Acknowledgements

We thank Ken Mayer for his involvement in the coordination of the study. Assay kits were kindly provided to Mayo Clinic for evaluations from Acon, Paramount (DeepBlue and Jinwofu), EcoTest (EcoTest Housing 2), and BTNX (EcoTest Housing 1). The work was supported by Herbert Irving Comprehensive Cancer Center in partnership with the Irving Institute for Clinical and Translational Research via a SARS-CoV-2 Research Pilot Grant, Columbia University School of Engineering and Applied Science via a Technology Innovations for Urban Living in the Face of COVID-19 Pilot Grant, and a gift from Bing Zhao.

## Author contributions

S.A., U.M., and S.K.S., conceptualized the initial project. S.A., J.M, S.C., and S.K.S. supervised the project. U.M. developed the object detection module. J.M., G.H., and S.C. developed the feature extractor and few-shot adaptation modules. D.I. and U.M. worked on cloud architecture and model hosting. A.Y. developed an image-labelling dashboard. D.C. developed the workflow to record kit parameters. S.A, J.M., U.M., S.C., and S.K.S. analyzed the data. K.M. and T.G. tested the assay kits with clinical samples and acquired smartphone images. R.S. helped label images for object detection. S.A., J.M., U.M., S.C., and S.K.S. wrote the manuscript.

## Author Declarations

Serum samples for the antibody tests were collected under Mayo Clinic IRB 20-004544 or shared by the Department of Laboratory Medicine at the University of Washington School of Medicine (Seattle, WA). The use of excess clinical specimens was reviewed by the Mayo Clinic Biospecimens Committee and an appropriate Material Transfer Agreement was drawn up to allow access to de-identified specimens from the University of Washington School of Medicine. The University of Washington IRB approved this work with a consent waiver. Nasopharyngeal swabs from Mayo Clinic Hospital patients were heat fixed and run for the antigen tests under Mayo Clinic IRB 20-010688.

