## Supplementary data for "Adaptable Automated Interpretation of Rapid Diagnostic Tests Using Few-Shot Learning"

### Supplementary Tables

| Kit name | Designation | No. of zones per kit | Kit aspect ratio | Membrane aspect ratio | Zone aspect ratio |
|---|---|---|---|---|---|
| EcoTest | Base | 3 | 0.29 | 0.21 | 3.23 |
| Flowflex | New | 2 | 0.28 | 0.22 | 2.94 |
| DeepBlue | New | 2 | 0.28 | 0.19 | 2.17 |
| Jinwofu | New | 2 | 0.30 | 0.21 | 3.03 |
| ACON IgG/IgM | New | 3 | 0.28 | 0.21 | 2.86 |
| EcoTest housing 2 | New | 3 | 0.53 | 0.21 | 3.13 |

**Supplementary Table 1. Assay kit parameters.** Summary of manually measured parameters for each of the assay kits.

| | Flowflex | | DeepBlue | | Jinwofu | | ACON IgG/IgM | | EcoTest housing 2 | |
|---|---|---|---|---|---|---|---|---|---|---|
| | Zone | Memb. | Zone | Memb. | Zone | Memb. | Zone | Memb. | Zone | Memb. |
| Positive class (train) | 149 | 50 | 125 | 25 | 146 | 50 | 182 | 63 | 20 | 5 |
| Negative class (train) | 51 | 50 | 75 | 75 | 54 | 50 | 118 | 37 | 10 | 5 |
| Positive class (evaluation) | 305 | 83 | 244 | 60 | 164 | 60 | 386 | 126 | 66 | 21 |
| Negative class (evaluation) | 143 | 141 | 124 | 124 | 44 | 44 | 193 | 67 | 6 | 3 |

**Supplementary Table 2. Dataset split of training and evaluation sets for 5 new kits.** The train and evaluation dataset sizes in terms of cropped zone images and membranes for the five new assay kits.

| Kit name | IOU score |
|---|---|
| Flowflex | 0.92 ± 0.009 |
| DeepBlue | 0.91 ± 0.012 |
| Jinwofu | 0.91 ± 0.079 |
| ACON IgG/IgM | 0.92 ± 0.012 |

**Supplementary Table 3. Mean IoU scores using bootstrapping.** The mean IOU scores for each of the new kit images, except for the EcoTest housing 2, was obtained by carrying out bootstrap sampling and obtaining three resamples. Each sample had ten randomly chosen images from a labelled pool of 30 images for training and the performance was evaluated on a fixed evaluation set of ten images. Due to a limitation on the available images only a single sampling was used for the EcoTest housing 2. Data is represented as mean ± standard deviation.

| Kit name | Zone accuracy (%) | Kit accuracy (%) |
|---|---|---|
| Flowflex | 99.6 ± 0.2 | 99.3 ± 0.3 |
| DeepBlue | 99.3 ± 0.2 | 98.6 ± 0.4 |
| Jinwofu | 99.6 ± 0.4 | 99.2 ± 0.7 |
| ACON IgG/IgM | 98.4 ± 0.5 | 95.3 ± 1.3 |

**Supplementary Table 4. Mean classification accuracy scores using bootstrapping.** The mean accuracy values were determined by carrying out 20 trials of bootstrap sampling with 10-shot adaptation (20 zone images) on the new kit's training dataset and evaluating the performance on the evaluation dataset. Data is represented as mean ± standard deviation.

**Supplementary Figures**

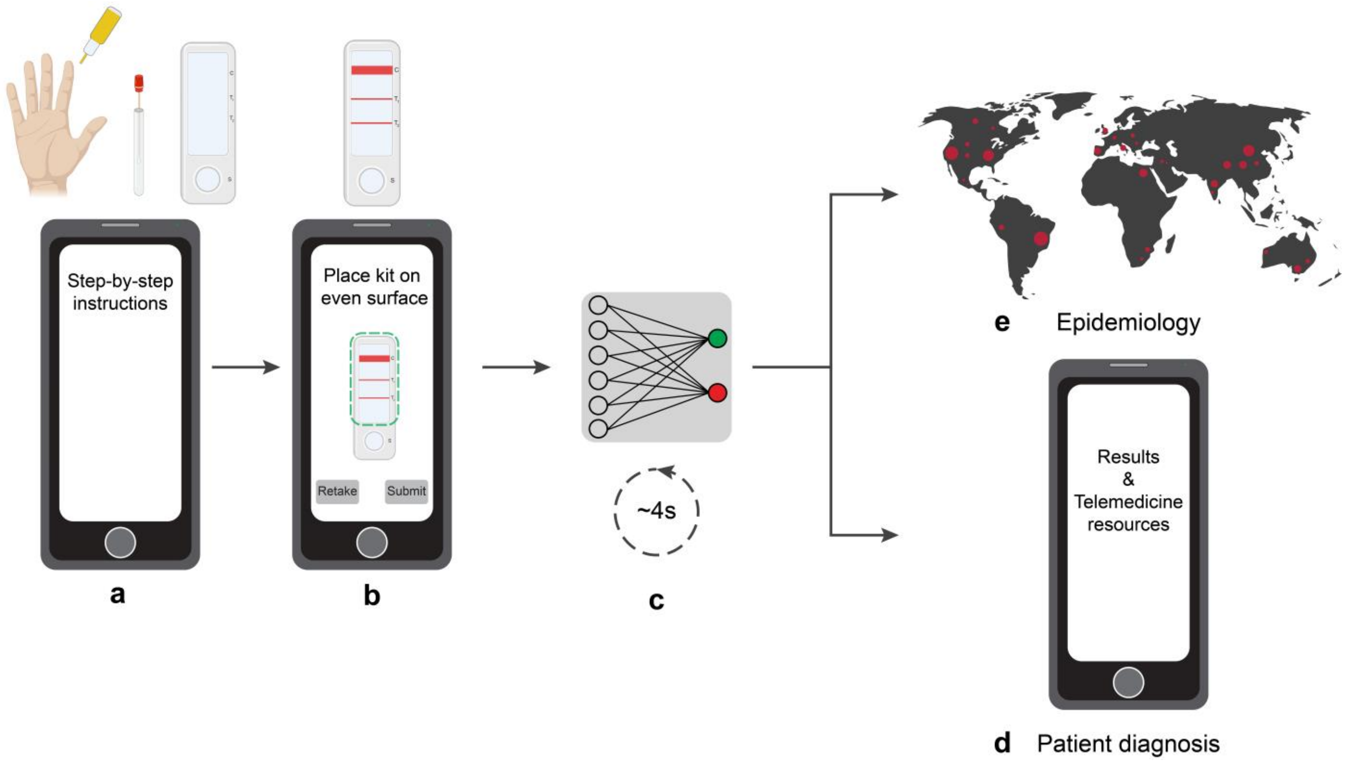


**Supplementary Figure 1. Potential role of AutoAdapt LFA within a broader digital platform for supporting rapid testing in decentralized settings**. **(a)** Step-by-step instructions to carry out the test. **(b)** App guided image capture. **(c)** Image preprocessing and deep-learning based classification. **(d)** Results displayed on user's phone and link to resources (sharing, saving, linkage-to-care). Automatically interpreted test results can also be used with telemedicine consultation. **(e)** Test results with demographic data stored on cloud server for real-time surveillance and modeling. (Created with BioRender.com)

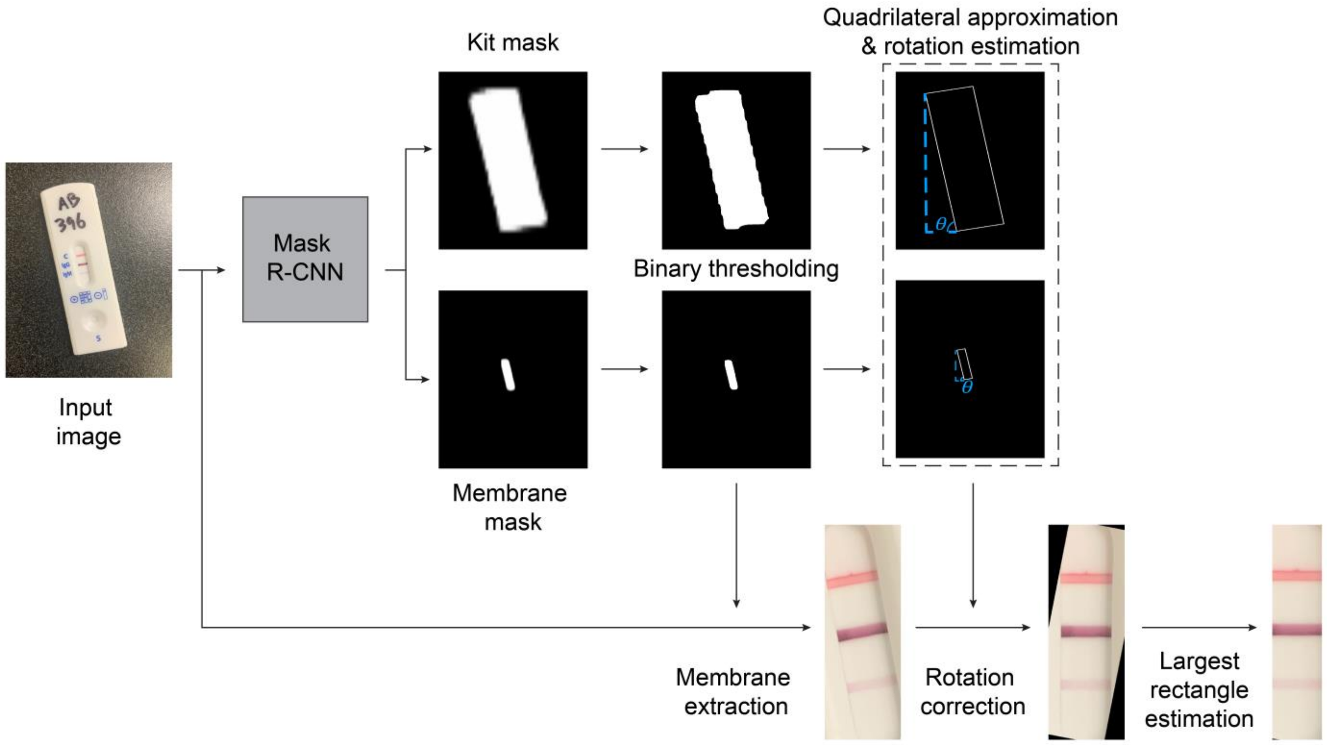


**Supplementary Figure 2. Workflow for membrane extraction and rotation correction.** The assay kit image is passed through the instance segmentation model Mask R-CNN. The kit and membrane segmentation masks are obtained and binarized. The kit or the membrane mask is used for quadrilateral approximation and rotation estimation in the image, and the membrane mask is used to extract the membrane from the input image. The estimated rotation value is used to correct the perspective of the extracted membrane. Largest rectangle estimation is carried out to remove the black pixels in the rotation-corrected membrane to retain only the relevant pixels

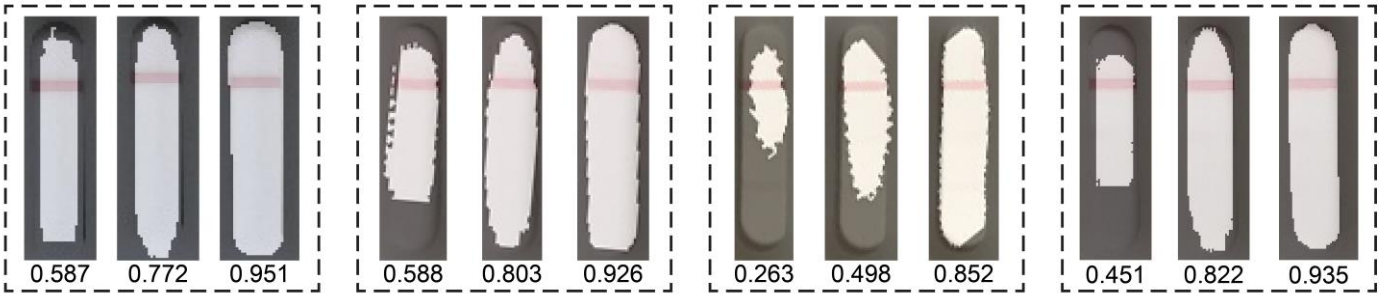


**Supplementary Figure 3. Illustration of different IoU scores for membrane segmentation.** Images shown are of the EcoTest (base kit) with the membrane segmentation mask overlaid on the zoomed-in image of the membrane. For each membrane image, segmentation masks corresponding to different IoU scores are shown with the score listed below the image.
